# Self-reported food choices before and during COVID-19 lockdown

**DOI:** 10.1101/2020.06.15.20131888

**Authors:** E. Siobhan Mitchell, Qiuchen Yang, Heather Behr, Laura Deluca, Paul Schaffer

**Affiliations:** Noom Inc., New York, NY, United States; Department of Integrative Health, Saybrook University, Oakland, CA, United States; Ferkauf Graduate School of Psychology, Yeshiva University, Bronx, NY, United States

**Keywords:** dietary intake, COVID-19, digital health, food groups, food logging

## Abstract

Stressful situations can cause changes in individual food choices, most notably, choices of highly rewarding foods that are high in fat or sugar. Few studies have examined how a population’s food choices change during a country-wide stress-inducing event such as the beginning of the COVID-19 stay-at-home orders in the United States. Food data from a digital behavior change weight loss program, which includes an interface for logging meals, were analyzed to assess self-reported food choices from March 5-March 11 (“pre-COVID”) and during the first week of the COVID-19 lockdown (March 12-March 18; “during-COVID”). The final sample consisted of 381,564 participants: 318,076 (83.4%) females, the majority who were aged 45-65 years (45.2%). Results indicate that self-reported servings of fresh fruit and vegetable intake decreased from pre-to during-COVID, while intake of red meat and starchy vegetables increased. More men than women increased their intake of red meat and processed meat. There was less overall change in fruit and vegetable consumption in participants aged 66 and older, compared to younger participants (aged 18-35). The percentage of older participants who reported lean meat and starchy vegetable intake increased, but these groups had a negligible change in younger subjects. More subjects aged 18-35 years reduced their intake of caffeine, desserts, lean meat and salads compared to older participants. No changes were observed in terms of snack or alcoholic beverage intake logged. In conclusion, this study of 381,564 US participants revealed that intake of particular food groups were altered during the first weeks of COVID lockdown.

## 1. Introduction

A growing body of evidence has suggested that stress not only affects our health through physiological processes but through changes in behavior such as food choice and total food intake (Torres & Nowson, 2007). Emotional eating has been defined as eating behaviors triggered by strong emotions and is initiated through stressful situations (Oliver & Wardle, 1999; Wallis & Hetherington, 2009). One commonly reported eating behavior in response to stressful situations is an inherent need to increase energy intake (Wansink et al., 2003); foods with higher sugar and fat content are more calorically dense and are frequently reported to be consumed during emotional or stressful periods (Oliver & Wardle, 1999).

While stress can influence food choice, food choice has shown to influence behavior (Gibson, 2006). One common way food can affect behavior is the change in mood that occurs after eating a meal (Gibson, 2006). Comfort foods are often chosen during times of emotional stress and are described as foods that induce a psychological change in state and promote comfort and pleasure for a person when consumed (van den Bos & de Ridder 2006). There is a wide array of comfort foods people choose to consume. It has been suggested that individual preference for food types is based on physiological needs (Geiker et al., 2017). Therefore, foods such as calorically dense snacks, rich desserts, chocolate, or ice cream may be comfort food for one person, while warm meals with meats or hearty soup may be comfort food for others. In an observational study, Wansink et al. (2003) reported differences in comfort food choices between age groups and gender. The youngest respondents and women preferred snack foods such as potato chips and ice cream as comfort foods. Older respondents and men were more inclined to choose meals as comfort food (Wansink et al., 2003).

Per (Geiker et al., 2017), the more relevant and threatening the stressor is perceived to be; the more stress one will experience. While countries issue lockdowns and travel bans due to the COVID-19 pandemic, many populations are having to cope with strong emotions and stress. The World Health Organization (WHO, 2020) has reported that uncertainty about the disease has prompted fear and anxiety as people are faced with new requirements for social distancing and growing economic wariness. In a crisis of this magnitude, it can be expected that many people will experience considerable changes to their eating behaviors and food choices as they cope with stress (Pietrobelli et al., 2020). The purpose of this study is to examine how a populations ‘ food choices changed during the COVID-19 stay-at-home orders imposed throughout the United States. It was expected that differences in food choices would be seen across age groups and gender.

## 2. Methods

### 2.1. Data collection

This is an observational, retrospective, cohort study. Data was collected via the behavioral change weight loss program, Noom. Noom is a mobile intervention which allows users to log their weight, meals and physical activity via a smartphone interface and also gives access to a virtual 1:1 behavior change coach, support group, and daily curriculum that includes diet-, exercise-, and psychology-based content (Jacobs et al. 2017). Noom users were initially self-referred and signed up for the program through the app store (iTunes/Google Play). Informed consent to participate in research is included during the sign-up phase of the program, in which users can choose to opt-out. This study, and informed consent, were approved by an external IRB.

Noom users ‘self-reported responses to the COVID-19 epidemic in the United States were assessed using an existing food database that had been previously developed by Noom. Data were collected from the pre-COVID time period (March 5-March 11) and during the first week of the COVID-19 lockdown (March 12-March 18). March 12 was designated as the first day after COVID-19 stay-at-home orders based on news articles advising people in several states to avoid non-essential shopping and to remain at home (Mervosh & Swales, 2020).

To be included in the study, users enrolled were (a) 18 years and older, and (b) living in the US. Users were excluded from the study if: (a) total usage of the program was less than three weeks (b) users did not enter food log data during both weeks of March 5-11 and March 12-18 and (c) users did not use the English language version of the program.

### 2.2. Statistical Analysis

Descriptive statistics were calculated for users ‘baseline characteristics and expressed in means, medians, and standard deviations for continuous variables and frequencies and percentages for categorical variables (Table 1). Frequencies and percentage of changes for food consumption data were compared before and after March 12 (Table 2, 3, 4).

**Table 1:**
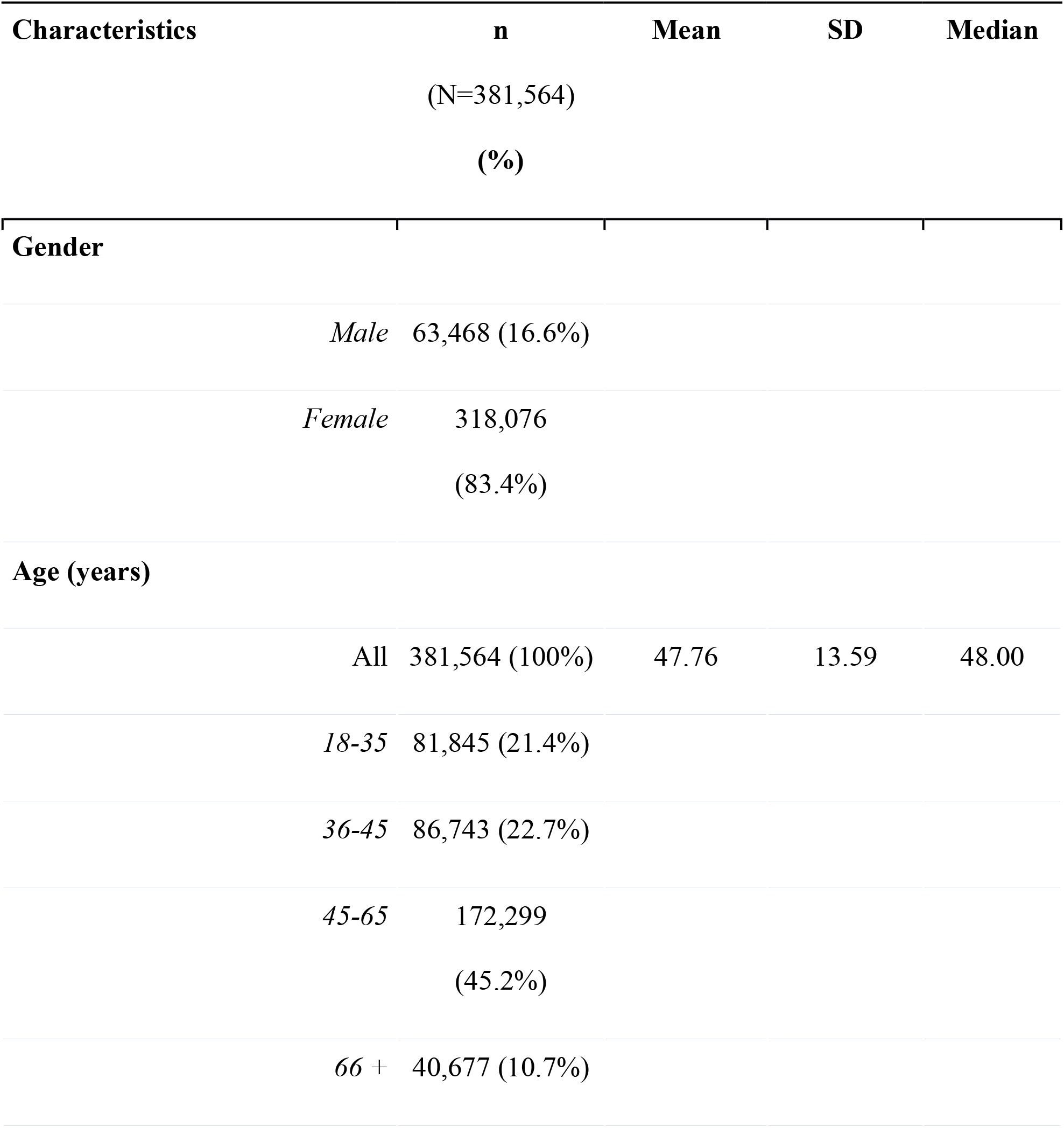
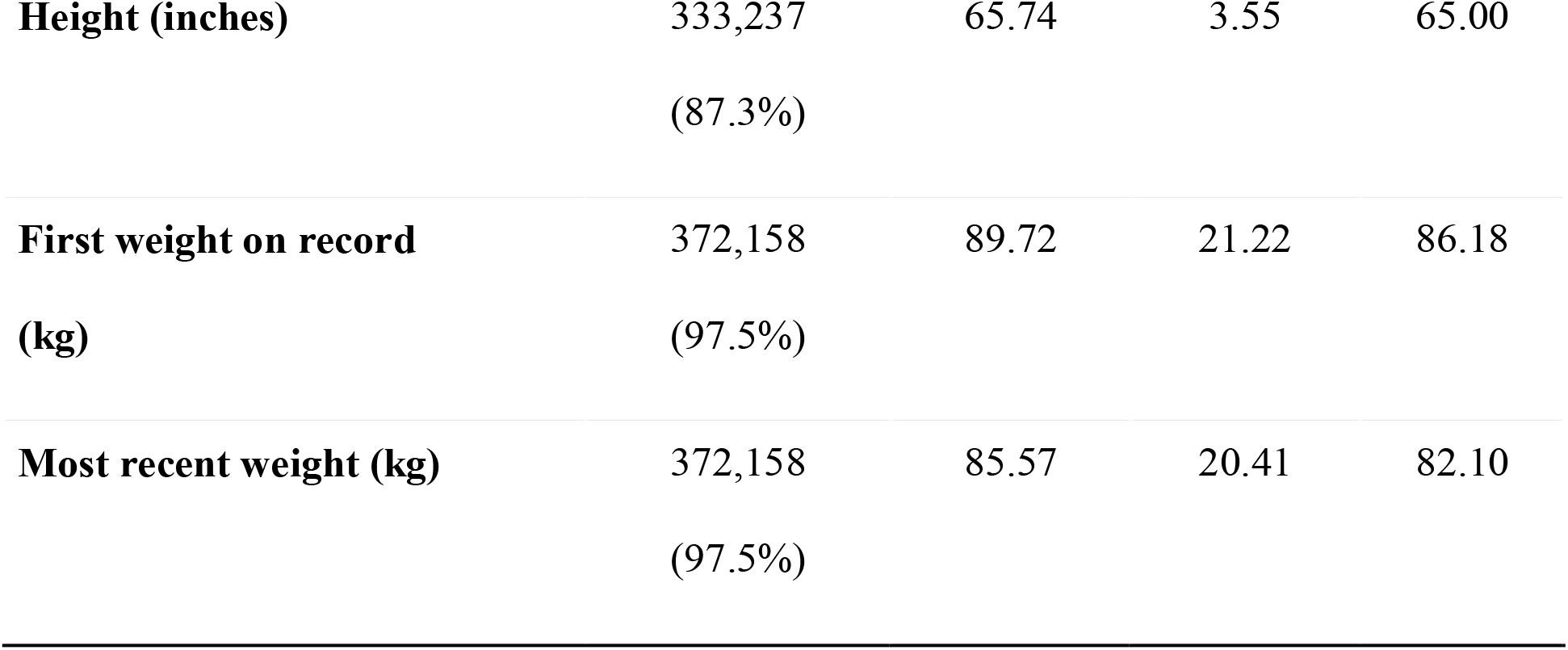
Demographic and weight information of participants who logged food during March 5-11 and March 12-18

**Table 2:**
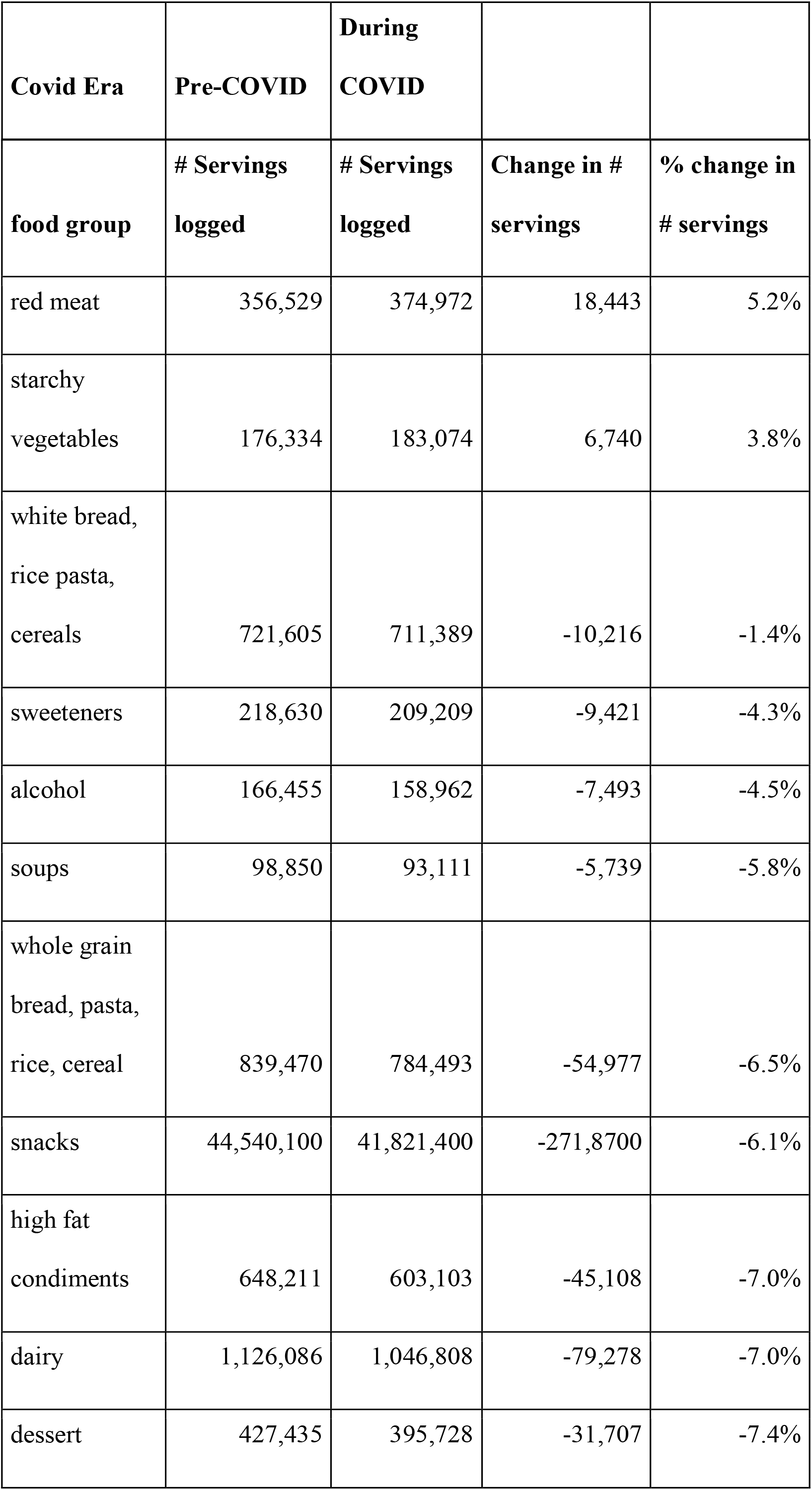
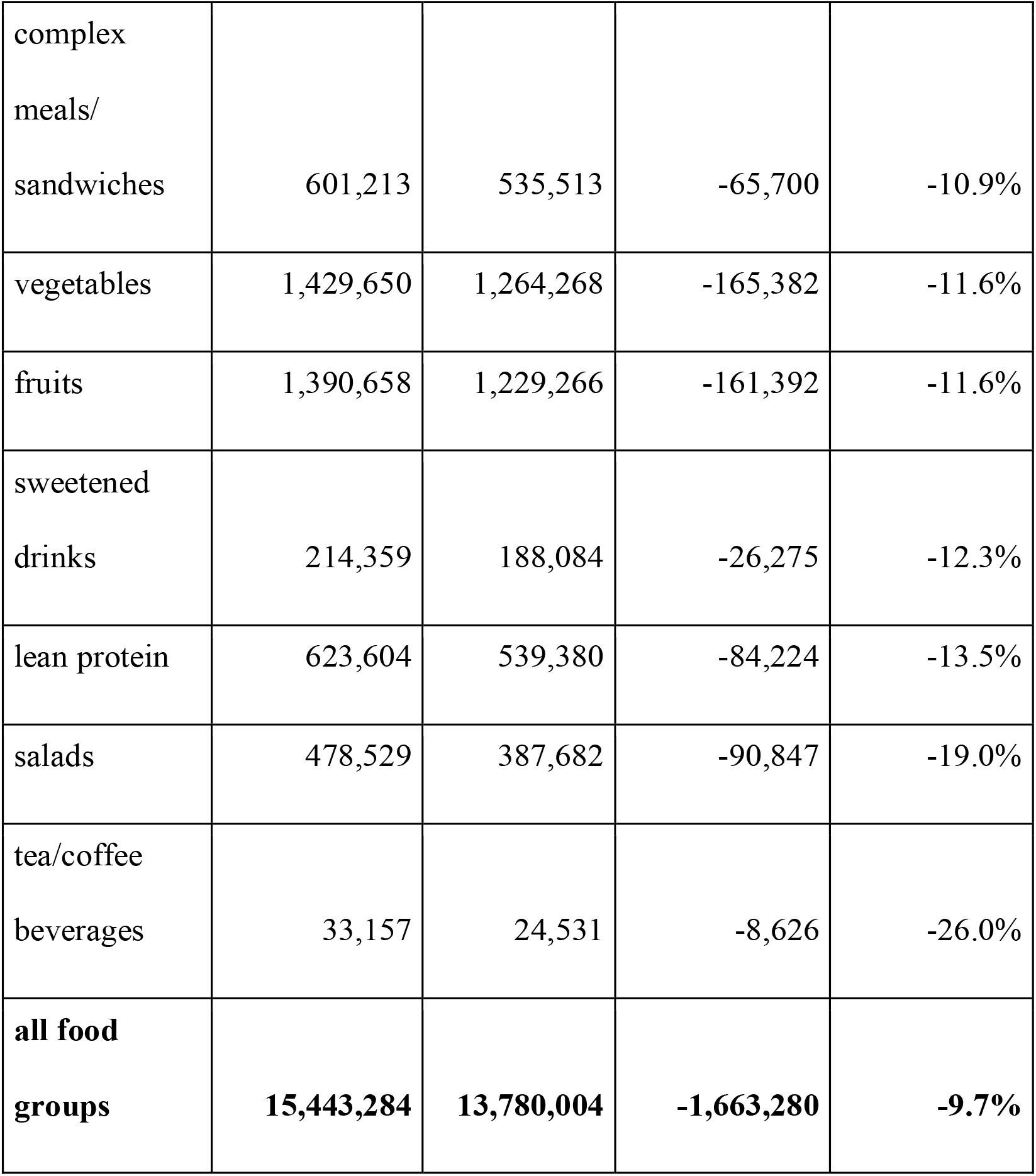
Number of servings according to the food group for March 5-11 (Before COVID) and March 12-18 (during COVID); change in the number of servings per week and % change in food group servings was also extracted.

**Table 3:**
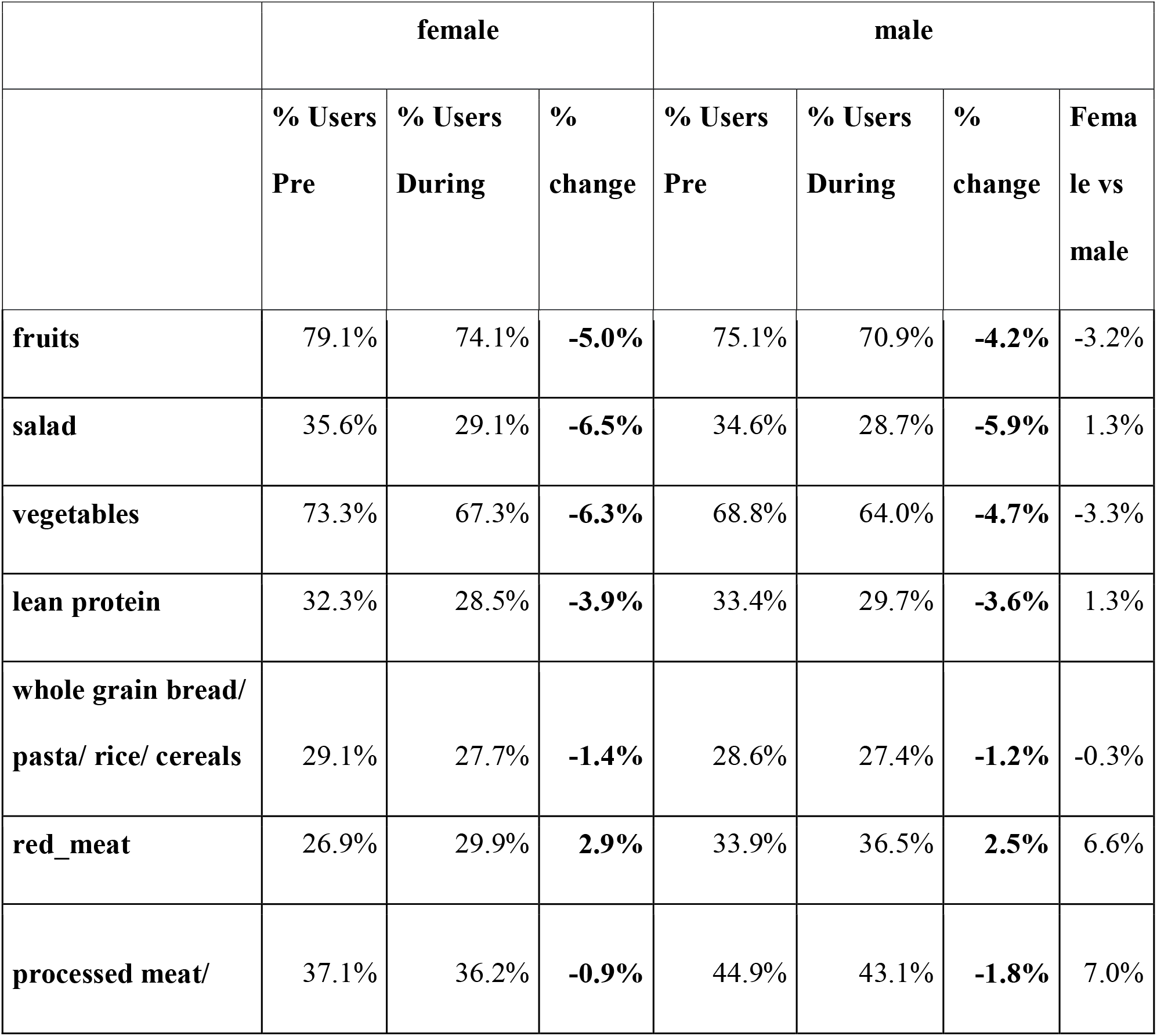
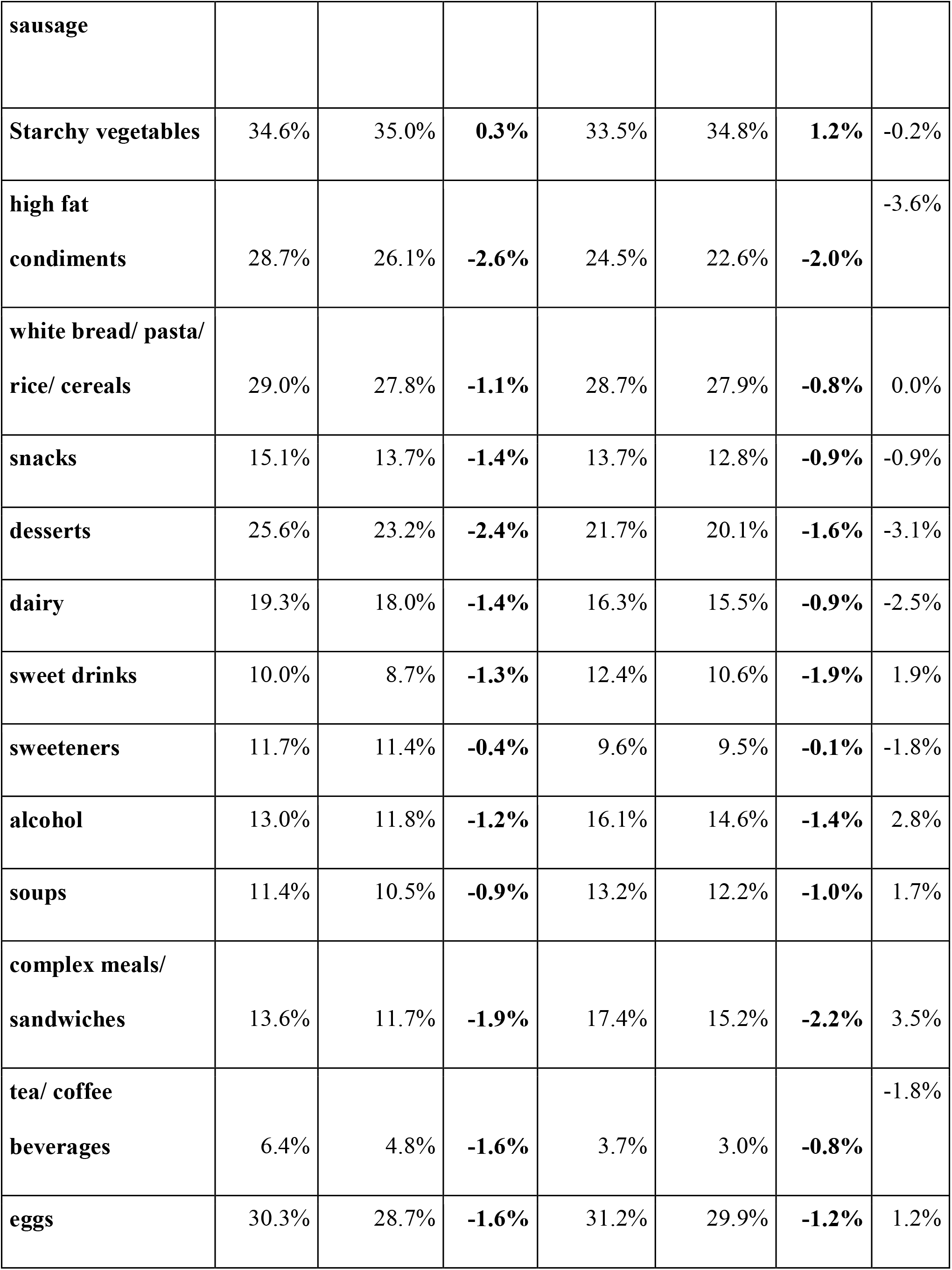
Percent of users reporting consumption of specific food groups according to gender (Pre: pre-COVID, March 5-11; During: During COVID, March 12-18)

|  | female |  |  | male |  |  |  |
| --- | --- | --- | --- | --- | --- | --- | --- |
|  | % Users<br>Pre | % Users<br>During | %<br>change | % Users<br>Pre | % Users<br>During | %<br>change | Fema<br>le vs<br>male |
| **fruits** | 79.1% | 74.1% | **-5.0%** | 75.1% | 70.9% | **-4.2%** | -3.2% |
| **salad** | 35.6% | 29.1% | **-6.5%** | 34.6% | 28.7% | **-5.9%** | 1.3% |
| **vegetables** | 73.3% | 67.3% | **-6.3%** | 68.8% | 64.0% | **-4.7%** | -3.3% |
| **lean protein** | 32.3% | 28.5% | **-3.9%** | 33.4% | 29.7% | **-3.6%** | 1.3% |
| **whole grain bread/**<br>**pasta/ rice/ cereals** | 29.1% | 27.7% | **-1.4%** | 28.6% | 27.4% | **-1.2%** | -0.3% |
| **red_meat** | 26.9% | 29.9% | **2.9%** | 33.9% | 36.5% | **2.5%** | 6.6% |
| **processed meat/** | 37.1% | 36.2% | **-0.9%** | 44.9% | 43.1% | **-1.8%** | 7.0% |
| **sausage** |  |  |  |  |  |  |  |
| **Starchy vegetables** | 34.6% | 35.0% | **0.3%** | 33.5% | 34.8% | **1.2%** | -0.2% |
| **high fat**<br>**condiments** | 28.7% | 26.1% | **-2.6%** | 24.5% | 22.6% | **-2.0%** | -3.6% |
| **white bread/ pasta/**<br>**rice/ cereals** | 29.0% | 27.8% | **-1.1%** | 28.7% | 27.9% | **-0.8%** | 0.0% |
| **snacks** | 15.1% | 13.7% | **-1.4%** | 13.7% | 12.8% | **-0.9%** | -0.9% |
| **desserts** | 25.6% | 23.2% | **-2.4%** | 21.7% | 20.1% | **-1.6%** | -3.1% |
| **dairy** | 19.3% | 18.0% | **-1.4%** | 16.3% | 15.5% | **-0.9%** | -2.5% |
| **sweet drinks** | 10.0% | 8.7% | **-1.3%** | 12.4% | 10.6% | **-1.9%** | 1.9% |
| **sweeteners** | 11.7% | 11.4% | **-0.4%** | 9.6% | 9.5% | **-0.1%** | -1.8% |
| **alcohol** | 13.0% | 11.8% | **-1.2%** | 16.1% | 14.6% | **-1.4%** | 2.8% |
| **soups** | 11.4% | 10.5% | **-0.9%** | 13.2% | 12.2% | **-1.0%** | 1.7% |
| **complex meals/**<br>**sandwiches** | 13.6% | 11.7% | **-1.9%** | 17.4% | 15.2% | **-2.2%** | 3.5% |
| **tea/ coffee**<br>**beverages** | 6.4% | 4.8% | **-1.6%** | 3.7% | 3.0% | **-0.8%** | -1.8% |
| **eggs** | 30.3% | 28.7% | **-1.6%** | 31.2% | 29.9% | **-1.2%** | 1.2% |

**Table 4.**
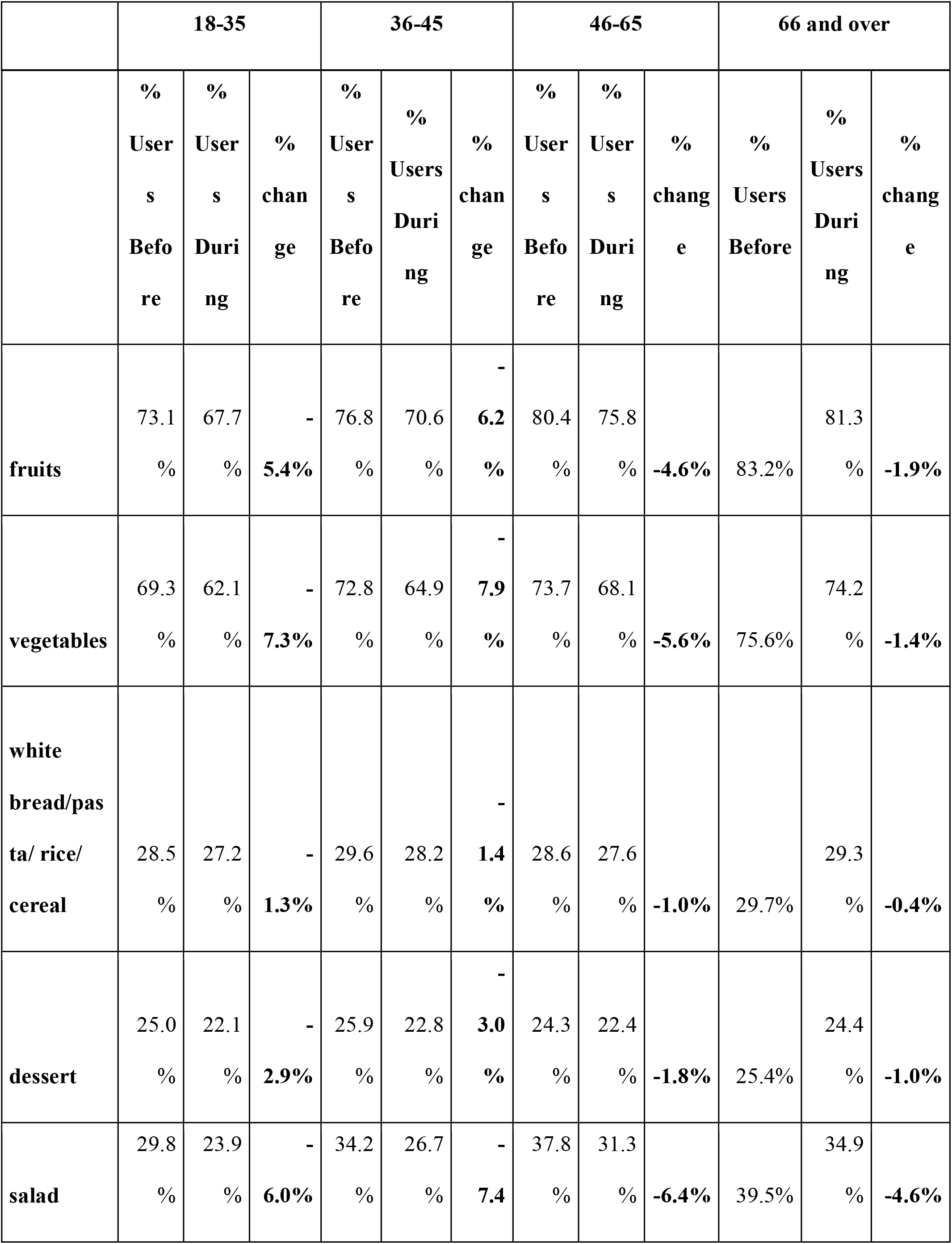
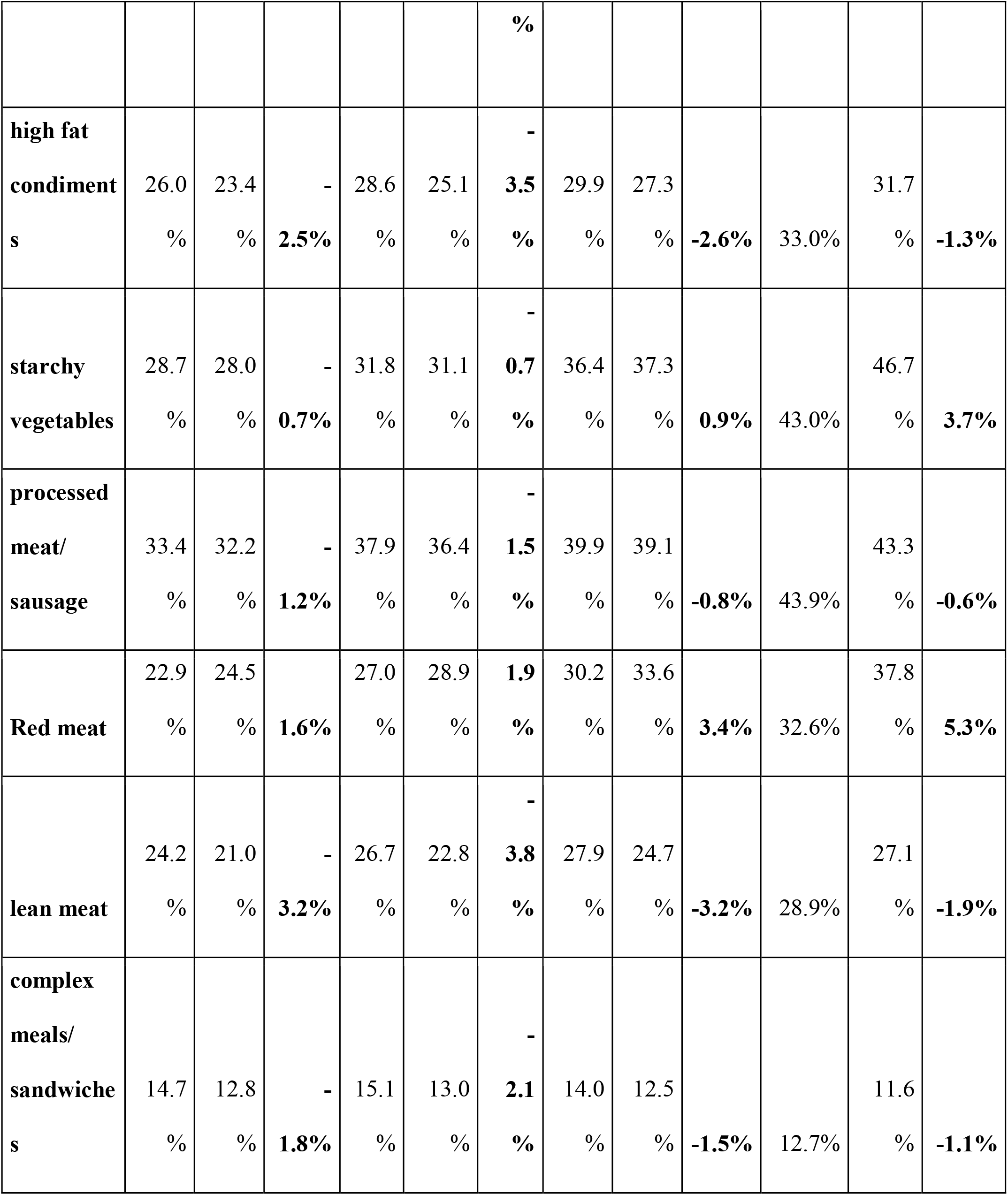
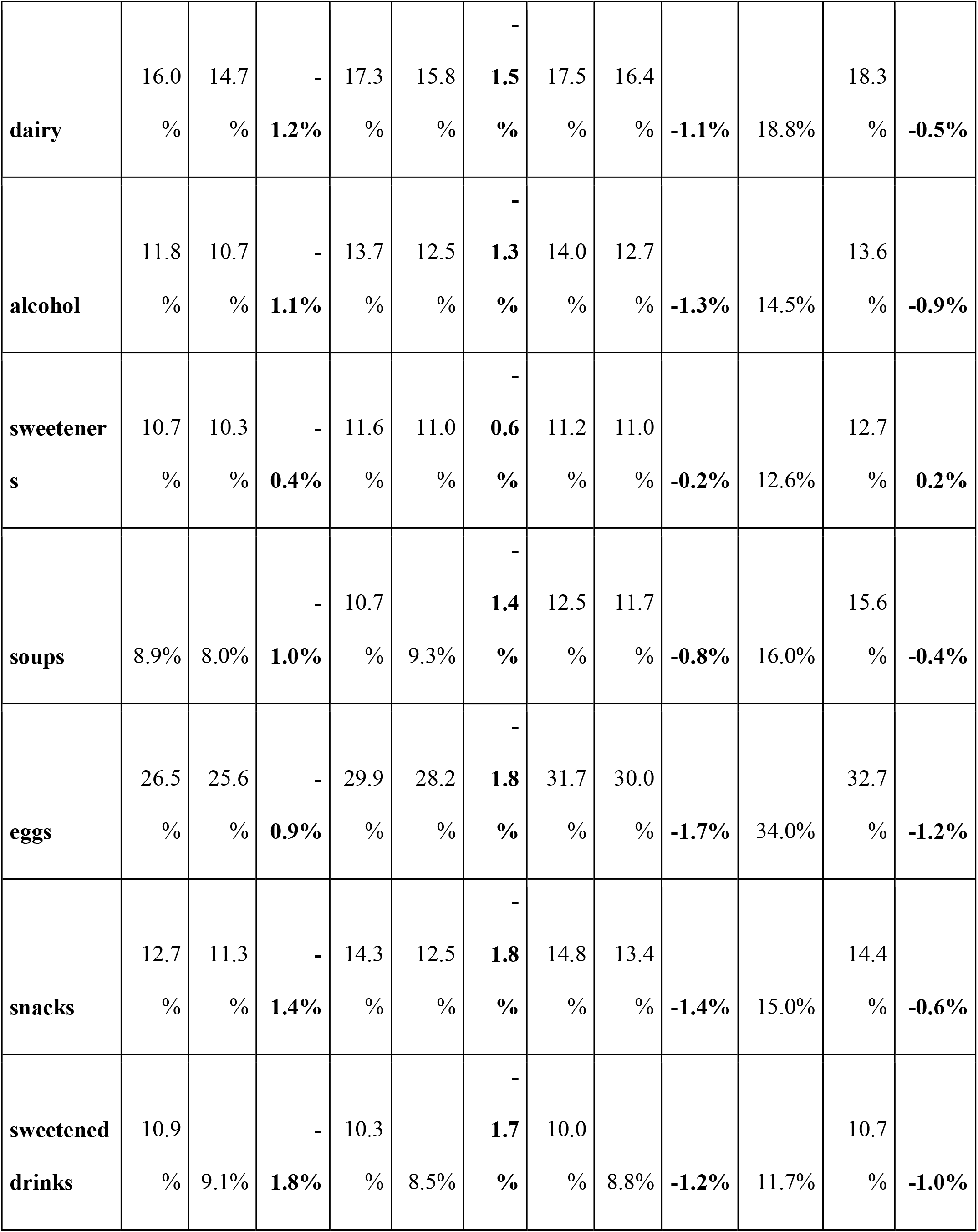
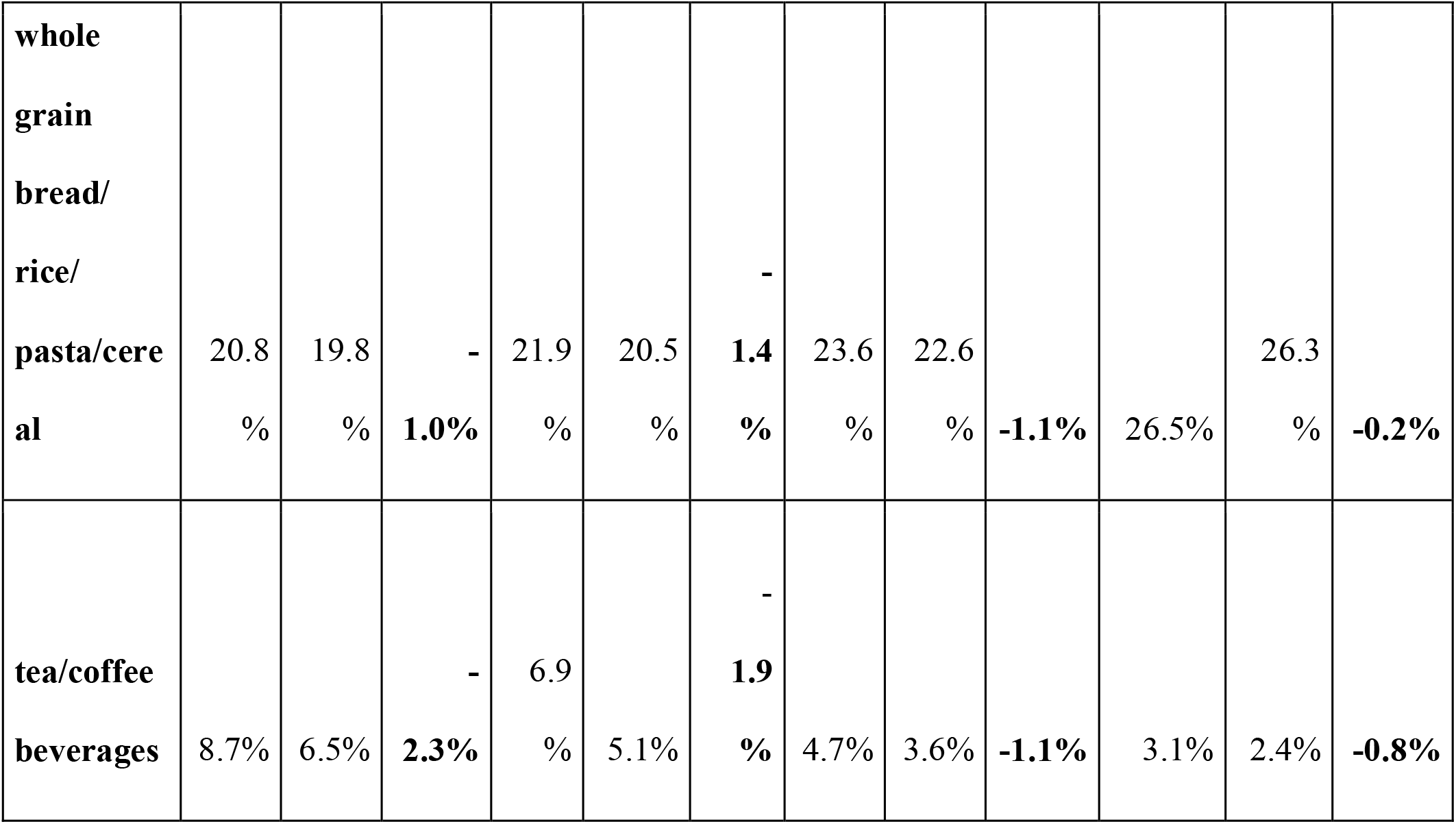
Percent of users reporting consumption of specific food groups according to age (Before: before COVID, March 5-11; During: During COVID, March 12-18)

### 2.3 Analysis of food groups

Analysis of food groups was extracted from food categorizations in the Noom food database, which contains thousands of food products available in the United States, and allows users to designate their preferred serving measurement unit: metric (g) or Imperial (oz) including units such as tablespoons, cups, bowls, plates, and typical serving units for specific foods (e.g., slices for apples or oranges). Products are categorized into food types reflecting the main component of the food or preparation of the food, which allows further compiling of food types into food groups (e.g., chips servings are compiled into a ‘snacks’ category, along with popcorn, nuts, pretzels, cheese puffs, and other salty snack foods). The food group’s complex meals/sandwiches include foods containing several food groups such as meat, grains, dairy, or vegetables that are prepared as a main meal. ‘Sweetened drinks’ include sugar-sweetened sodas, as well as diet soda, juice, and juice drinks. Since the Noom program encourages healthy eating, foods containing whole grains, such as whole wheat flour, are given a different designation than foods made from white flour, such as regular pasta or white bread. Thus ‘whole grains ‘designates cereals primarily made with whole-grain ingredients, such as brown rice, corn, or whole wheat. ‘High-fat condiments ‘include butter, oils, high-fat dressings, sauces, and cream. ‘Red meat ‘includes fatty cuts of meat or preparations such as hamburgers, steaks, or lamb or pork chops. ‘Lean protein ‘ includes lean cuts of pork and beef, chicken, seafood, soy-based foods, and protein shakes. ‘Processed meats and sausage ‘includes fatty sausage, bacon, and other highly processed meats.

## 3. Results

Of the 381,564 participants who met inclusion criteria, 318,076 (83.4%) were females, and the mean age of the sample was 47.76 (SD=13.59) (Table 1). The majority of users included in our population were aged 45-65 years old (45.2%). The average self-reported first weight on record for participants is 89.72 kg (SD=21.2), and the most recent average weight is 85.57 kg (SD= 20.4).

To assess changes in dietary patterns, data from Noom users who logged meals during the week of March 5-March 11 (Pre-COVID) and during the week of March 12-18 (during COVID) was analyzed according to food grouping. Users’ overall logging of food group servings decreased by 9% in terms of actual items of food logged (Table 2). Results indicate that users increased intake of red meat (5%) and starchy vegetables (potatoes, corn, peas, and winter squash, 4%). Lean meat, salads, and caffeinated drinks show large decreases in logging (i.e., those food groups exhibiting a larger overall drop than the general decrease in food group logging). Moderate decreases in vegetables, fruit, and sweetened drinks were also observed during this time period.

When food group data were analyzed according to self-identified gender, several differences in overall consumption patterns between males and females were observed. The largest differences in self-reported intake were seen in the greater percentage of men versus women reporting meat food groups: red meat (6.6% difference) and processed meats/sausage (7% difference), and also in the complex meals/sandwiches group (3.5% difference) (Table 3). A higher percentage of female users reported intake of fruits (3.2%) and dessert (3.1%) than male users. Both men and women had an increase in the percentage of users reporting red meat (men: 2.5%; women: 2.9%) during the first week of COVID stay-at-home mandates. Overall, more food groups decreased in terms of the percentage of users reporting consumption: Notably fruits (men: −4.2%; women:- 5.0%), salad (men:-5.9%; women: −6.5%) vegetables (men: −4.7%; women:-6.3%), and lean protein (men:-3.6%; women:-3.9%) dropped in terms of users reporting intake.

When data was analyzed among age groups, other patterns emerged on how older versus younger users changed their diet during the initiation of COVID stay-at-home policies (Table 4). The self-reported intake of food groups changed much less in users aged 66 and older than users aged 18-35 years from the week of March 5 to the week of March 12. There were decreases in the percentage of younger users reporting consumption of fruits and vegetables (−5.4% and - 7.3%, respectively) during the week of March 12, yet negligible change in the percentage of older users ‘consumption of fruits and vegetables. Conversely, the increase in users aged 66 and over who consumed red meat was substantially higher (5.3%) than younger age groups. A similar pattern was also seen in the percentage of older users consuming starchy vegetables (+3.7% for users 66 and older vs −0.7% for users aged 18-35 years). Meanwhile, the percentage of younger users consuming lean meat rose and slightly decreased in users 66 and older.

All groups showed a drop in salad intake (−6%, −7.4%, −6.4%, and −4.6% for age groups 18-35, 36-45, 46-65, and 66 and over, respectively). While the percentage of users consuming alcohol did not change according to age, caffeinated drinks such as tea and coffee decreased in the proportion of users aged 18-35 years (−2.3%) but only marginally decreased in users aged 35 and older.

## 4. Discussion

Our study sought to explore Noom users ‘food choices during the COVID-19 stay-at-home orders imposed throughout the United States. Results showed that fruit and vegetable consumption, especially salads, decreased during the initial week of US COVID stay-at-home mandates, while red meat, processed meat and starchy vegetable consumption increased. A higher percentage of men and older participants (66 years and older) reported red meat and starchy vegetable intake during March 12-18 vs. March 5-11.

Few studies have examined food choices after a stressful event. To our knowledge, three studies reported food choice changes after a natural disaster. Women living in Christchurch, New Zealand, engaged in emotional eating (more snacking and less fruit and vegetable consumption) after a 7.1 Richter scale earthquake (Kuijer & Boyce 2012). Another study assessed food group intake in evacuees after the 2011 Fukushima Earthquake in eastern Japan and reported that diet quality worsened in those reporting higher stress, i.e., less intake of fruits and vegetables, meat, soy, and dairy (Uemura et al., 2016). A follow-up study in the same population also demonstrated that living in non-home environments also contributed to poor diet quality (Zhang et al., 2017).

Interestingly, in the current study, subjects are living at home but may be unable or unwilling to move about freely, which also limits access to fresh food. Due to the nature of the study population, it is not possible to determine if the change in diet was due to stress or decreased mobility. Also, we are unable to determine if the overall diversity of food decreased or if users simply did not log food as frequently during the first week of COVID lockdown.

The desire for high-calorie food is explained as a tendency towards short term reward after prolonged stress (van den Bos & de Ridder, 2006). The population we surveyed are taking part in a behavior change program, and so it could be construed that many users struggle with emotional eating, and indeed, stress-induced eating is observed more in overweight subjects than normal-weight subjects (Corning & Viducich 2020). However, since the Noom program is not specifically designed to measure stress and mood, we are not able to measure associations of comfort food eating to stress levels. Nor are we able to consider the diet patterns of the users according to their level of lockdown. Nevertheless, the current set of data agrees with the news coverage of decreased fruit and vegetable purchasing in favor of more shelf-stable foods and red meat (Creswell 2020). Thus, it appears that food choices during this period may be reflecting desires to stock up and consume core food groups such as red meat and starchy vegetables.

Indeed, similar results have been reported in a study examining children’s eating behavior during COVID lockdown (Pietrobelli et al. 2020). Meanwhile, the percentage of users consuming alcohol, regardless of age or gender, did not differ during the two consecutive weeks, while caffeine consumption decreased in the youngest age range (18-35 years), which may be due to less opportunity or desire to drink caffeinated beverages at home than in a workplace.

There are several limitations to this study. One limitation is that all data are self-reported meal logging, which is susceptible to flawed recall, omissions, or lapses in logging. While some studies have indicated a reasonable correlation of digital food logging data to more traditional methods, results can vary according to the platform used (Rangan et al. 2016; Boushey et al. 2017). The participant data is drawn from a population enrolled in a weight loss program and may not reflect the food choices of those without weight concerns. Additionally, the participant pool is skewed more towards women (83.4%) than men (16.6%). Lastly, changes in COVID-related eating behavior depend on the extent of lockdown in different regions of the US and may not have occurred at the same time periods. However, the large number of participants included in the study (n=381,564) adds robustness to the findings.

In conclusion, this observational study explored how food choices changed during the initiation of US COVID stay-at-home orders according to age and gender in a population engaged in weight loss. Further research is needed to understand the factors that influenced food choices and how food choices are modified in similar and dissimilar populations.

## Data Availability

The data used for this study is stored according to HIPAA regulations, de-personalized data will be provided upon request

